# Factors associated with bacterial sexually transmitted infections amongst people of South Asian ethnicity in England

**DOI:** 10.1101/2023.05.17.23290085

**Authors:** R Dhairyawan, A Shah, JV Bailey, H Mohammed

## Abstract

**Objectives:** Despite being the largest ethnic minority group in England, South Asians have historically had low levels of utilisation of sexual health services and sexually transmitted infection (STI) diagnoses, though recent data suggests this may be changing. This study aimed to investigate factors associated with a bacterial STI diagnosis amongst South Asians attending SHS in England.

**Methods:** Using data from the GUMCAD STI Surveillance system, a descriptive analysis of South Asians attending sexual health services in England in 2019 was carried out. Factors associated with a bacterial STI diagnosis were examined using univariate and multivariable logistic regression models adjusted for age, Asian ethnic subgroup, HIV status, patient region of residence and Index of Multiple Deprivation quintile. Analyses were stratified by gender and sexual orientation (heterosexual male vs. gay, bisexual, and other men who have sex with men [GBMSM] vs. women of any sexual orientation). Crude and adjusted associations were derived using binary logistic regression.

**Results:** There were 121,842 attendances by South Asians to SHS in England in 2019. Compared to heterosexual South Asian men, GBMSM had a higher odds of being diagnosed with a bacterial STI (aOR 2.32, 95% CI 2.19-2.44) and South Asian women had a lower odds (aOR 0.83, 95% CI 0.78-0.87). For women and heterosexual South Asian men, a diagnosis was associated with younger age, being of any other Asian background other than Bangladeshi, Indian or Pakistani and not being HIV positive. For heterosexual South Asian men, there was an association with increasing socioeconomic deprivation. For GBMSM, a bacterial STI diagnosis was associated with known HIV positive status and living in London.

**Conclusions:** People of South Asian ethnicity in England are heterogenous with regards to their sexual health needs, which should be explored further through focused research and policy.

**Key Messages:**

- What is already known on this topic: Historically South Asians in England have had low rates of sexually transmitted infections and sexual health service utilisation. Despite South Asians being the largest ethnic minority group in England, there has been a paucity of research investigating their sexual health needs.
- What this study adds: To our knowledge, this is the first national epidemiological study focusing on people of South Asian ethnicity accessing sexual health services in England, finding variation in factors associated with bacterial STI diagnosis, particularly when stratified by gender and sexual orientation.
- How this study might affect research, practice or policy: Our results reflect the heterogeneity of South Asian communities in England with regards to sexual health need, and we suggest that policy makers and researchers should prioritise this under-served group to improve their sexual health outcomes.

## Introduction

Globally, people living in South Asia bear a high burden of sexually transmitted infections (STIs)^1^. In England however, people of South Asian ethnicity have historically had low levels of STI diagnoses and utilisation of sexual health services (SHS)^[2-4]^. South Asians living with HIV in England are also often diagnosed late, an important risk factor for HIV-related morbidity and mortality, suggesting missed opportunities to access HIV testing^5,6^. Research has found that barriers to South Asians accessing SHS in the UK include a lack of risk perception, poor knowledge about sexual health, stigma and shame, and SHS that are inconvenient or provide a poor service-user experience^[7-17]^.

Despite being the largest ethnic minority group in the UK, there has been a paucity of epidemiological research studies focusing on South Asian communities and their sexual health in the UK. This has been attributed to low STI rates compared to other ethnic groups, and concerns from researchers about the possible cultural sensitivities of the topic^15^.

Whilst people of Black Caribbean ethnicity had the highest rates of STIs in England in 2019, the largest proportional increase in all new STI diagnoses since 2018 was in people of Asian ethnicity (16%, from 15,168 to 17,522 total diagnoses)^4^. Among people of Asian ethnicity, this increase was mainly gonorrhoea (36%) and chlamydia (27%) in gay, bisexual and other men who have sex with men (GBMSM)^4^. The reasons for this increase are unclear, but they may include shifts in sexual health care seeking or sexual risk behaviour. There continues to be community interest in discussing sexual health in the media^18^.

This study aims to investigate factors associated with a bacterial STI diagnosis amongst South Asians attending SHS in England using the national STI surveillance dataset.

## Methods

### Study Design

This is an observational study using data on attendances at SHS in England, reported to the UK Health Security Agency as part of the GUMCAD STI Surveillance System (GUMCAD)^19^. This includes data on demographics, service attendances, STI tests and diagnoses. GUMCAD is a pseudonymised, depersonalised dataset of all attendances at SHS in England. All services, tests and diagnoses are coded by healthcare practitioners in keeping with surveillance reporting specifications. To avoid double counting of diagnoses, only one diagnosis is retained within a 6-week episode^19^. GUMCAD does not include direct identifiers such as name or date of birth, so unique individuals are identified using a clinic-specific patient identification code.

The study sample included all people aged ≥15 years of a South Asian ethnic group who attended an SHS in 2019 for an STI-related reason (an attendance where they were tested for or diagnosed with an STI). This excluded people attending SHS solely for contraception. 2019 was chosen as the most recent year before the COVID-19 pandemic, which caused considerable disruption to service delivery^20^.

### Key variables

South Asian ethnicity was defined as those who self-reported as Asian or Asian British ethnicity (Indian, Bangladeshi, Pakistani, Any Other Asian Background). We also included people for whom ethnicity was not specified, but who were born in a South Asian country (India, Pakistan, Bangladesh, Sri Lanka, Bhutan, Nepal and the Maldive Islands)^21^.

The outcome variable was binary – being diagnosed with a bacterial STI or not, with bacterial STIs defined as chlamydia, gonorrhoea and infectious syphilis (primary, secondary or early latent stages).

Co-variates with *a priori* important clinical and public health relevance were chosen. These were gender and sexual orientation (heterosexual men, GBMSM, heterosexual women, women who have sex with women [WSW]), age group, born in the UK vs not born in the UK, and HIV status (known positive vs negative/unknown). Also, attendees’ region of residence (London, Midlands and East of England, North of England, South of England), the urban vs rural composition of the area and Index of Multiple Deprivation (IMD) quintile of residence (derived from service users’ Lower Layer Super Output Area of residence), with 1 containing the most, and 5 containing the least deprived areas.

### Statistical analysis

#### Descriptive analyses were carried out to describe the characteristics of people attending SHS

Pearson’s Chi Square tests were carried out to look for associations between the main independent variable (South Asian ethnic subgroup), outcome (bacterial STI diagnosis) and co-variates. This identified potential confounders. However, due the size of the GUMCAD dataset, analyses can be over-powered increasing the risk of a Type 1 error. Therefore, confounders were only included in analysis if known *a priori* and of public health or clinical relevance. These were sexual and gender orientation, age, ethnic subgroup, IMD quintile, HIV status and patient region of residence.

Univariate and multivariable logistic regression models were performed to determine factors associated with a bacterial STI diagnosis, reported with unadjusted odds ratios (OR) and adjusted odds ratios (aOR) with 95% confidence intervals (CI). Models were adjusted using *a priori* confounders and covariates with unadjusted associations with a p-value <0.05. Models were created using a forward stepwise approach, adding a variable at a time. Model fit was assessed using Akaike Information Criterion tests. Univariate and multivariable models were stratified by gender and sexual orientation, as these have been found *a priori* to have important clinical and public health relevance. As numbers of WSW were small, heterosexual women and WSW were combined into one category. The strata used for analysis were: heterosexual men, GBMSM and women of any sexual orientation.

Sensitivity analyses were carried out to look at the Any Other Asian Background category in more detail. This included univariate and multivariable logistic regression models to determine factors associated with a bacterial STI diagnosis, using the same confounders used for the sample of South Asians as a whole. Country and region of birth were also determined.

#### Analyses was carried out using Stata v17.0

This study was granted ethical approval by the UKHSA Research and Public Health Practice Ethics and Governance Group and was reviewed by members of community interest group South Asian HIV Advisory Resource (SAHAR).

## Results

There were 121,842 STI-related attendances by South Asians to SHS in England in 2019. Table 1 describes the characteristics of the South Asian attendees. 40.83% were heterosexual women with 32.24% heterosexual men, 17.20% GBMSM, 0.35% WSW and 9.39% unknown gender and sexual orientation. The majority (62.60%) were aged 24-44 years. The largest South Asian ethnic subgroup was Indian (35.02%). Almost the same proportion were born in the UK (43.39%) as born outside (41.16%). More than half (56.42%) lived in regions that were most deprived (IMD quintile ranks 1 and 2). Most lived in predominantly urban areas (90.91%) with almost a half living in London (47.78%). A minority (4.25%) were known to be living with HIV. There were 7.54% diagnosed with one or more bacterial STI.

**Table 1.** Characteristics of South Asians attending sexual health services for STI related reasons in England in 2019

| Variable | Category | N=121842 | % |
| --- | --- | --- | --- |
| Gender and sexual orientation | Heterosexual male | 39281 | 32.24 |
|  | GBMSM | 20952 | 17.20 |
|  | Heterosexual female | 49744 | 40.83 |
|  | WSW | 430 | 0.35 |
|  | Not specified | 11435 | 9.39 |
| Age group (years) | 15-19 | 7578 | 6.22 |
|  | 20-24 | 24835 | 20.38 |
|  | 25-34 | 49076 | 40.28 |
|  | 35-44 | 27290 | 22.40 |
|  | 45-64 | 11760 | 9.65 |
|  | 65+ | 885 | 0.73 |
|  | Not known | 418 | 0.34 |
| Ethnic sub-group | Indian | 42665 | 35.02 |
|  | Pakistani | 28311 | 23.24 |
|  | Bangladeshi | 14270 | 11.71 |
|  | Any Other Asian background | 36596 | 30.04 |
| Born in UK | Yes | 52871 | 43.39 |
|  | No | 50146 | 41.16 |
|  | Not known | 18825 | 15.45 |
| IMD quintile rank | 1 (most deprived) | 32313 | 26.52 |
|  | 2 | 36436 | 29.90 |
|  | 3 | 24084 | 19.77 |
|  | 4 | 15022 | 12.33 |
|  | 5 (least deprived) | 9796 | 8.04 |
|  | Not known | 4191 | 3.44 |
| HIV status | Known HIV positive | 5182 | 4.25 |
|  | HIV negative or not known | 116660 | 95.75 |
| Region of residence | London | 58216 | 47.78 |
|  | Midlands and East of England | 28923 | 23.74 |
|  | North of England | 17493 | 14.36 |
|  | South of England | 14175 | 11.63 |
|  | Not known | 3035 | 2.49 |
| Rural Vs Urban | Predominantly Rural | 2142 | 1.76 |
|  | Predominantly Urban | 110765 | 90.91 |
|  | Urban and rural | 3762 | 3.09 |
|  | Not known | 5173 | 4.24 |
| Bacterial STI diagnosis | Yes | 9190 | 7.54 |
|  | No | 112652 | 92.46 |
| Chlamydia diagnosis | Yes | 6319 | 5.19 |
|  | No | 115523 | 94.81 |
| Gonorrhoea diagnosis | Yes | 3182 | 2.61 |
|  | No | 118660 | 97.39 |
| Syphilis diagnosis | Yes | 408 | 0.33 |
|  | No | 121434 | 99.67 |

For the logistic regression analysis, there were 105,919 attendances (87%) with complete data for all the variables of interest. Table 2 shows that on the unadjusted analysis, compared to heterosexual South Asian men, GBMSM had 2.28 higher odds of being diagnosed with a bacterial STI, which was similar after adjustment for confounders (aOR 2.32, 95% CI 2.19-2.44). Compared to heterosexual men, South Asian women had 0.91 lower odds of bacterial STI which reduced further on the adjusted analysis (aOR 0.83, 95% CI 0.78-0.87). The Any Other Asian ethnic subgroup were most likely to be diagnosed with a bacterial STI, with Indians least likely (aOR 0.77, 95%CI 0.72-0.81) compared to them.

**Table 2.** Factors associated with being diagnosed with a bacterial STI amongst South Asians in 2019. Univariate and multivariable analyses.

| Variable |  | Unadjusted |  |  | Adjusted* |  |  |
| --- | --- | --- | --- | --- | --- | --- | --- |
|  |  | OR | 95% CI | P-value | OR | 95% CI | P-value |
| Gender and sexual orientation | Heterosexual male | 1.00 | - | - | 1.00 | - | - |
|  | GBMSM | 2.28 | 2.16-2.42 | <0.001 | 2.32 | 2.19-2.44 | <0.001 |
|  | Female | 0.91 | 0.86-0.96 | 0.001 | 0.82 | 0.78-0.87 | <0.001 |
| Age group (years) | 15-19 | 1.00 | - | - | 1.00 | - | - |
|  | 20-24 | 0.79 | 0.72-0.86 | <0.001 | 0.75 | 0.69-0.82 | <0.001 |
|  | 25-34 | 0.63 | 0.58-0.68 | <0.001 | 0.53 | 0.49-0.58 | <0.001 |
|  | 35-44 | 0.52 | 0.47-0.57 | <0.001 | 0.43 | 0.39-0.47 | <0.001 |
|  | 45-64 | 0.43 | 0.38-0.48 | <0.001 | 0.36 | 0.32-0.40 | <0.001 |
|  | 65+ | 0.11 | 0.06-0.22 | <0.001 | 0.10 | 0.05-0.20 | <0.001 |
| Asian Ethnic subgroup | Any other Asian background | 1.00 | - | - | 1.00 | - | - |
|  | Indian | 0.69 | 0.66-0.73 | <0.001 | 0.77 | 0.72-0.81 | <0.001 |
|  | Pakistani | 0.88 | 0.83-0.93 | <0.001 | 0.90 | 0.85-0.96 | 0.001 |
|  | Bangladeshi | 0.84 | 0.77-0.91 | <0.001 | 0.89 | 0.82-0.97 | 0.006 |
| HIV status | HIV negative or not known | 1.00 | - | - | 1.00 | - | - |
|  | Known HIV positive | 1.24 | 1.12-1.38 | <0.001 | 1.04 | 0.93-1.16 | 0.503 |
| IMD quintile | 1 (most deprived) | 1.00 | - | - | 1.00 | - | - |
|  | 2 | 0.94 | 0.89-0.99 | 0.040 | 0.92 | 0.87-0.98 | 0.005 |
|  | 3 | 0.89 | 0.83-0.94 | <0.001 | 0.88 | 0.82-0.94 | <0.001 |
|  | 4 | 0.83 | 0.77-0.90 | <0.001 | 0.83 | 0.77-0.90 | <0.001 |
|  | 5 (least deprived) | 0.78 | 0.72-0.86 | <0.001 | 0.81 | 0.74-0.90 | <0.001 |
| Region of residence | London | 1.00 | - | - | 1.00 | - | - |
|  | Midlands and East of England | 0.99 | 0.94-1.05 | 0.802 | 1.06 | 1.00-1.12 | 0.037 |
|  | North of England | 0.95 | 0.89-1.01 | 0.110 | 0.97 | 0.90-1.04 | 0.347 |
|  | South of England | 0.96 | 0.89-1.03 | 0.246 | 0.99 | 0.92-1.07 | 0.894 |

\*Adjusted for sexual and gender orientation, age, ethnic subgroup, IMD quintile, HIV status, patient region of residence

Table 3 shows that among South Asian women and heterosexual men, the odds of a bacterial STI decreased with increasing age, and this relationship persisted after adjustment for confounders. Compared to the Any Other Asian Background group, the odds of a STI diagnosis was lowest for Indian heterosexual men in the univariate and the multivariable analyes (aOR 0.68, 95% CI 0.61-0.75) and women (aOR 0.73, 95% CI 0.66-0.80). Living with HIV was associated with a 73% lower odds of a bacterial STI for heterosexual men, compared to those HIV negative or of unknown status. On adjustment this was reduced to 62% (aOR 0.38, 95% CI 0.23-0.61). Women living with HIV had a 90% lower odds of a bacterial STI on the univariate analysis and 84% on the multivariable analysis (aOR 0.16, 95% CI 0.07-0.37) compared to women of HIV negative or of unknown status. Among heterosexual men, there was increasing odds of a bacterial STI when living in a more deprived area, but this was slightly attenuated after adjustment for confounders. Heterosexual men living outside of London had increased odds of being diagnosed with a bacterial STI, an association that persisted after adjustment.

**Table 3.**
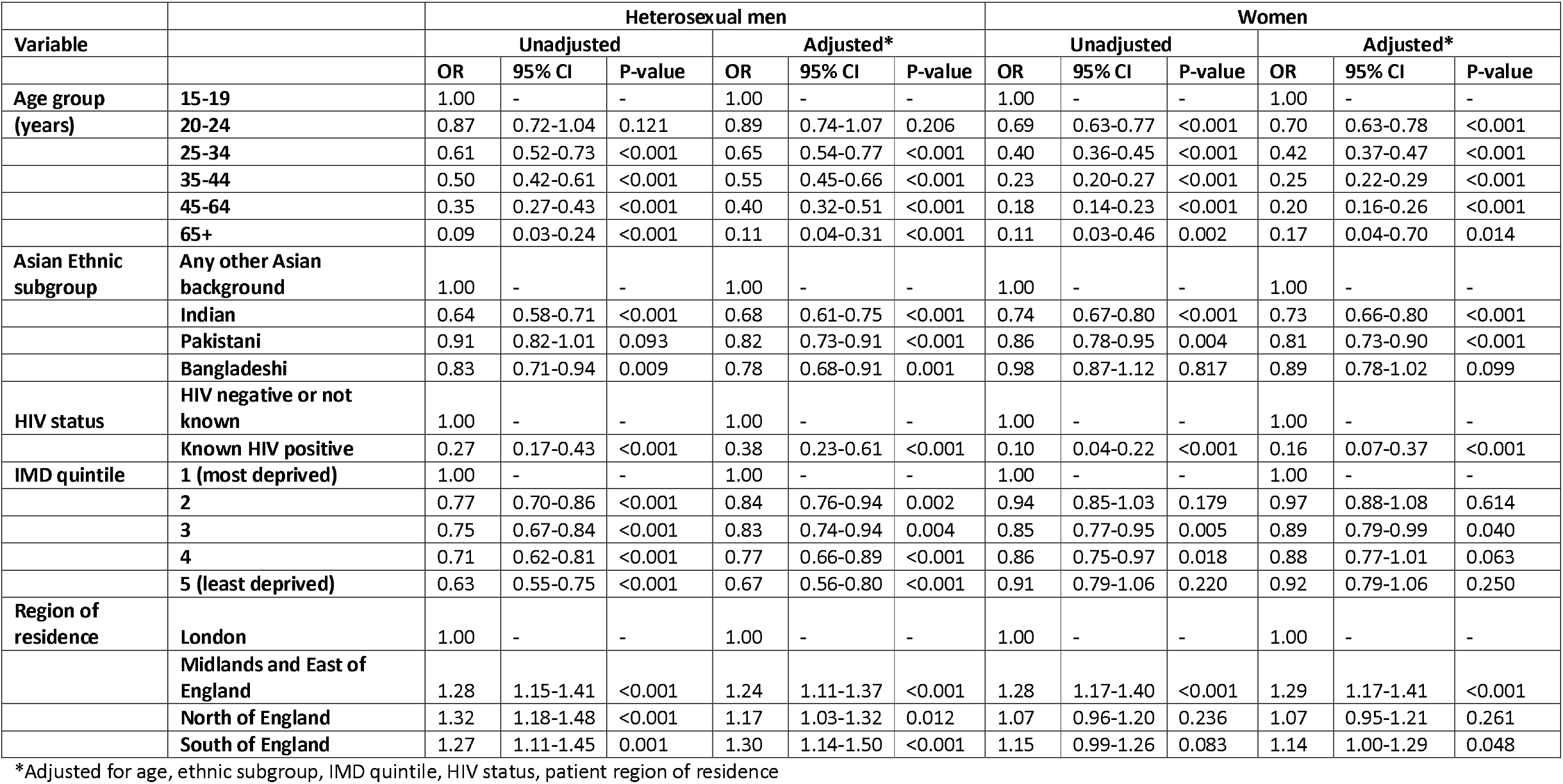
Factors associated with being diagnosed with a bacterial STI amongst South Asians in 2019. Univariate and multivariable analyses stratified by gender and sexual orientation.

Finally, Table 4 shows that for South Asian GBMSM, age was not associated with a bacterial STI diagnosis, apart from the oldest age group (>65 years) having a lower odds on the unadjusted analysis (OR 0.28, 95% CI 0.12-0.38) and adjusted analyses (aOR 0.27, 95% CI 0.08-0.87) compared to the youngest group (15-19 year olds). There was also no association with Asian ethnic subgroup except for Indian GBMSM who had 14% lower odds than the Any Other Asian Background group (aOR 0.86, 95% CI 0.78-0.95). GBMSM living with HIV were more likely to be diagnosed with a bacterial STI on the univariate analysis (OR 1.14, 95% CI 1.01-1.28) and this association was strengthened after adjustment for confounders (aOR 1.18, 95% CI 1.04-1.33). There was no statistically significant association between IMD quintile of deprivation and bacterial STIs except for the least deprived IMD rank compared to the most deprived, but this was no longer associated after adjustment for confounders. There was a strong association between South Asian GBMSM living in London being more likely to be diagnosed with a bacterial STI compared to those living outside on the univariate and multivariable analyses.

**Table 4.** Factors associated with being diagnosed with a bacterial STI amongst South Asian GBMSM in 2019.

|  |  | GBMSM |  |  |  |  |  |
| --- | --- | --- | --- | --- | --- | --- | --- |
| Variable |  | Unadjusted |  |  | Adjusted* |  |  |
|  |  | OR | 95% CI | P-value | OR | 95% CI | P-value |
| Age group<br>(years) | 15-19 | 1.00 | - | - | 1.00 | - | - |
|  | 20-24 | 1.13 | 0.88-1.46 | 0.342 | 1.11 | 0.85-1.42 | 0.461 |
|  | 25-34 | 1.17 | 0.92-1.48 | 0.206 | 1.11 | 0.87-1.40 | 0.492 |
|  | 35-44 | 1.09 | 0.86-1.39 | 0.477 | 1.02 | 0.80-1.30 | 0.878 |
|  | 45-64 | 1.03 | 0.79-1.34 | 0.809 | 0.96 | 0.74-1.26 | 0.784 |
|  | 65+ | 0.28 | 0.12-0.38 | 0.036 | 0.27 | 0.08-0.87 | 0.029 |
| Asian Ethnic subgroup | Any other Asian background | 1.00 | - | - | 1.00 | - | - |
|  | Indian | 0.73 | 0.67-0.80 | <0.001 | 0.86 | 0.78-0.95 | 0.003 |
|  | Pakistani | 0.85 | 0.77-0.94 | 0.002 | 1.06 | 0.95-1.18 | 0.322 |
|  | Bangladeshi | 0.98 | 0.87-1.12 | 0.803 | 0.97 | 0.82-1.14 | 0.684 |
| HIV Status | HIV negative or not known | 1.00 | - | - | 1.00 | - | - |
|  | Known HIV positive | 1.14 | 1.01-1.28 | 0.030 | 1.18 | 1.04-1.33 | 0.009 |
| IMD quintile | 1 (most deprived) | 1.00 | - | - | 1.00 | - | - |
|  | 2 | 1.02 | 0.91-1.13 | 0.717 | 0.96 | 0.86-1.07 | 0.477 |
|  | 3 | 0.98 | 0.86-1.09 | 0.670 | 0.93 | 0.82-1.05 | 0.252 |
|  | 4 | 0.89 | 0.77-1.02 | 0.083 | 0.86 | 0.76-1.01 | 0.060 |
|  | 5 (least deprived) | 0.83 | 0.70-0.99 | 0.031 | 0.86 | 0.72-1.03 | 0.103 |
| Region of residence | London | 1.00 | - | - | 1.00 | - | - |
|  | Midlands and East of England | 0.78 | 0.70-0.87 | <0.001 | 0.78 | 0.70-0.88 | <0.001 |
|  | North of England | 0.81 | 0.71-0.92 | 0.002 | 0.75 | 0.65-0.87 | <0.001 |
|  | South of England | 0.78 | 0.67-0.89 | <0.001 | 0.79 | 0.68-0.91 | 0.001 |

\*Adjusted for age, ethnic subgroup, IMD quintile, HIV status, patient region of residence

Focusing on the Any Other Asian Background group (n=36,596), we found that attendees recorded countries of birth from all world regions. Table 1 in the Supplementary Material shows the world regions of birth. The sensitivity analysis conducted showed similar results for this group with the South Asian sample as a whole, with regards to factors associated with a bacterial STI analysis.

## Discussion

To our knowledge, this is the first national epidemiological study to focus on the sexual health of people of South Asian ethnicity in England using national surveillance data. Our results demonstrate variation in factors associated with bacterial STI diagnosis amongst South Asians by gender and sexual orientation – highlighting the extent of heterogeneity within these populations. South Asian gay and bisexual and other men who have sex with men are twice as likely, and women one-fifth less likely, to be diagnosed with a bacterial STI than heterosexual South Asian men. After stratifying by gender and sexual orientation, there were similarities for women and heterosexual men, among whom bacterial STI diagnosis was associated with younger age, being of Any Other Asian Background ethnic subgroup and not being HIV positive. For heterosexual South Asian men, there was an association with increased socioeconomic deprivation and living outside of London. For GBMSM, living in London or living with diagnosed HIV were associated with a bacterial STI diagnosis.

Our results are consistent with national trends showing high rates of bacterial STIs amongst GBMSM in England, which have risen from 2012-2019^22^. This has been attributed to increased testing, but also sexual behaviours such as multiple condomless anal intercourse partners^23^. A survey found that racially minoritised GBMSM including Asians were more likely to report condomless anal intercourse with more than one non-steady partner than White GBMSM^24^. Racially minoritised GBMSM experience discrimination in multiple settings which may reduce their ability to practice safer sex. South Asian GBMSM in the UK may experience homophobia within their ethnic communities, leading them to conceal their sexuality,^25^ and also may experience marginalisation in gay communities. Racially minoritised GBMSM in the UK report that experiencing racism within ‘gay spaces’ reduces the impact of sexual health promotion in these spaces^26^.

Similar to national trends, there was a strong association between younger age and a bacterial STI diagnosis, for South Asian women and heterosexual men. Young people are more likely to have higher rates of partner change^27^. Specific to British South Asian communities, a shift in cultural attitudes to sex in younger generations means that sex before marriage, and with partners from another ethnic groups, has become more common over time^11^.

There is a well-established link between socio-economic deprivation and poor health outcomes such as STIs in England^4^, which has been demonstrated in this study for heterosexual South Asian men. In 2019, Asians were the ethnic group most likely to live in the most deprived 10% of neighbourhoods in England, but this mostly comprised Pakistani and Bangladeshi communities rather than Indian communities^28^. This may partially explain why in our study Indians had the lowest odds of a bacterial STI diagnosis.

Consistent with national trends in England since 2009,^29^ South Asian GBMSM living with HIV had increasing rates of bacterial STIs compared to HIV negative GBMSM. However, for women and heterosexual men, living with HIV was associated with reduced odds of a bacterial STI. There may be several reasons for this including reduced knowledge about Undetectable=Untransmittable, fewer sexual partners and intersectional stigmas particularly for women, which should be further investigated. It may also reflect different patterns in healthcare utilisation among people living with HIV, with GBMSM perhaps attending SHS more regularly for sexual health screens than heterosexuals.

South Asian GBMSM living in London were more likely to be diagnosed with a bacterial STI, consistent with national trends showing London has twice the rates of new STIs than any other region in England, and most of these are seen in GBMSM^30^.

There are strengths to this study. As it uses data from a mandatory surveillance dataset in England, it is likely to be nationally representative of all South Asians attending SHS. The sample size is also large, increasing the power of the study to find statistically significant differences.

There are limitations which should be considered when interpreting the results and drawing inferences. Firstly, classification of ethnicity has inherent limitations which may bias the results. How people identify themselves may not match the categories provided, and the category ‘Any Other Asian Background’ contains people born in regions outside of those we have defined as being ‘South Asian’ so we have captured people who may not have South Asian heritage. GUMCAD also measures attendances and cannot identify whether individuals have attended several different SHS, leading to some double counting should people attend multiple SHS over the same episode of care. Some South Asians may seek sexual health advice and STI testing outside of specialist SHS, such as at their general practice, and this activity is not captured in GUMCAD. We do not know if these people may have different demographic characteristics to those accessing SHS. We did not report separate findings for WSW, as their numbers were too small for meaningful analysis and so were combined with heterosexual women as a category.

Finally, bacterial STIs are not the only markers of sexual health and wellbeing. We therefore cannot make strong inferences about a group’s sexual health from these results. Whilst the results may be applicable to other countries with similar health systems in Western Europe, we cannot say that they generalisable to all South Asians globally.

To our knowledge, this is the first study that has used national STI surveillance data to focus on people of South Asian ethnicity in England and therefore provides novel insights. We found that the Any Other Asian background ethnic subgroup were most likely to be diagnosed with bacterial STI suggesting a high level of sexual health need. We need to better understand who comprises this ethnic subgroup, as by looking at region of birth, it may not only include people of South Asian origin.

We hope that the findings of our analysis may be of interest to clinicians, researchers, policy makers and people from South Asian communities, providing the basis for future work in this area.

With regards to surveillance, this could include further analyses by disaggregate ethnic groups, to closely monitor changing patterns that suggest increased sexual health needs. Qualitative researchers should explore sexual behaviours, sexual networks and attitudes to sexual health. Studies must go beyond individual behaviours to explore structural issues that impact on sexual health and access to care, such as the impact of racism, sexism, Islamophobia, homophobia and other forms of oppression. A large proportion of our sample were born abroad and the impact of citizenship status, time in the UK, whether a first or subsequent generation migrant and level of English literacy on sexual health should be explored. It’s likely that these may affect knowledge of how to navigate healthcare systems, and attitudes to sex, sexual orientation and sexual health. Studies should also look at sexual health in the broader sense going beyond STIs to for example, experiences of sexual violence, sexual wellbeing and pleasure.

Compared to other ethnic groups in England, national trends show that South Asians have the lowest low levels of STI diagnoses and utilisation of sexual health services (SHS)^[2-4]^. However, low use of SHS does not mean that South Asians have low sexual health needs. Policy makers and clinicians should explore ways to measure unmet need and respond accordingly. People of South Asian ethnicity have reported experiencing multiple barriers to accessing SHS^[7-17]^, so work must be done to make services more acceptable and welcoming. This is particularly timely with changes to SHS due to the COVID-19 pandemic and an increase in digital access, which may represent both opportunities and challenges.

Understanding how to improve community knowledge of sexual health and access to SHS is vital. Interventional studies could provide an evidence base for policy makers to explore what works to improve the sexual health of South Asians. This could include looking at relationship and sex education at school.

In conclusion, our analysis demonstrates heterogeneity within South Asian populations in England with regards to bacterial STI diagnoses. This should be explored further, as low utilisation of SHS may not mean low need, but could indicate high unmet sexual health need.

## Data Availability

All data produced in the present work are contained in the manuscript

