## Supplemental Table 1 for "Factors associated with bacterial sexually transmitted infections amongst people of South Asian ethnicity in England"

**Supplementary Material**

Table 1. Top 5 World regions of birth for the Any Other Asian Background ethnic subgroup.

| **World region of birth** | **N** | **%** |
| --- | --- | --- |
| Other* | 15,900 | 43.45 |
| UK | 8,814 | 24.08 |
| South Asia | 4,863 | 13.29 |
| European Union | 783 | 2.14 |
| Sub Saharan Africa | 603 | 1.65 |

*Other includes Middle East, Australasia, East Asia
